# Estimation of the basic reproduction number, average incubation time, asymptomatic infection rate, and case fatality rate for COVID-19: Meta-analysis and sensitivity analysis

**DOI:** 10.1101/2020.04.28.20083758

**Authors:** Wenqing He, Grace Y. Yi, Yayuan Zhu

## Abstract

The coronavirus disease 2019 (COVID-19) has been found to be caused by the severe acute respiratory syndrome coronavirus 2 (SARS-CoV-2). However, comprehensive knowledge of COVID-19 remains incomplete and many important features are still unknown. This manuscripts conduct a meta-analysis and a sensitivity study to answer the questions: What is the basic reproduction number? How long is the incubation time of the disease on average? What portion of infections are asymptomatic? And ultimately, what is the case fatality rate? Our studies estimate the basic reproduction number to be 3.15 with the 95% interval (2.41, 3.90), the average incubation time to be 5.08 days with the 95% confidence interval (4.77, 5.39) (in day), the asymptomatic infection rate to be 46% with the 95% confidence interval (18.48%, 73.60%), and the case fatality rate to be 2.72% with 95% confidence interval (1.29%, 4.16%) where asymptomatic infections are accounted for.

## 1 Introduction

Since the first case of the coronavirus disease 2019 (COVID-19) was found in Wuhan, P. R. China in December 2019, the disease has rapidly spread in the city of Wuhan, then to Hubei Province, China, and subsequently, across the world [1]. On March 11, 2020, the World Health Organization (WHO) declared COVID-19 to be a pandemic. The swift spread of the virus is largely attributed to its stealth transmissions for which infected patients may be asymptomatic or exhibit only flu-like symptoms in the early stage. Undetected transmissions present a remarkable challenge for the containment of the virus and pose an appalling threat to the public health. To understand the drastically negative impacts of COVID-19 on the public health, it is urgent to investigate key features pertinent to the disease: How severe is the transmission? How long is the incubation time of the disease on average? How many infections are asymptomatic? And ultimately, what is the case fatality rate?

To evaluate the severity of the virus spread, it is useful to estimate the *basic reproduction number* (denoted *R*_0_), defined as the average number of cases generated by an infected individual in a population where everyone is susceptible to infection. If the basic reproduction number *R*_0_ is larger than 1, the outbreak is regarded as self-sustaining unless control measures are implemented to mitigate the transmission [5]. Defined as the time from the moment of exposure to the virus until signs and symptoms of COVID-19 appear, the *incubation time* of a COVID-19 infected patient provides a useful measure for the disease development. Knowing the average incubation time of the COVID-19 patients is important for disease surveillance. To determine how deadly the COVID-19 is, it is fundamental to evaluate the *case fatality rate* which is calculated as the ratio of the number of deaths from COVID-19 to the number of infected cases.

Since the outbreak of the disease, a large body of research on COVID-19 has been done and many articles have been published in scientific journals or shared on platforms such as bioRxir and medRxir. Simulations of the epidemic have been published under various assumptions to delineate hidden transmissions of the virus [5]. While estimates of those important quantities have been reported in the literature, those results are quite different and vary considerably from study to study. There has been a lack of consensus of those estimates because of serious concerns on the heterogeneity among the studies. Different studies have been carried out on different patients under different conditions, and different authors may make different model assumptions. Interpreting the available findings must be coupled with the associated features of the studies.

Moreover, COVID-19 data contain substantial errors in that the number of confirmed cases is considerably under-reported, which is attributed to two primary reasons. Insufficient test kits do not allow every potential patient with COVID-19-like symptoms to be tested, and there has been a good portion of asymptomatic COVID-19 carriers who would never be tested and counted as confirmed cases. It is useful to understand the *asymptomatic infection rate*, defined as the ratio of the number of asymptomatic infections to the number of all infected cases.

To address these issues, we carry out a meta-analysis to synthesize the reported estimates of the basic reproduction number, the average incubation time, and the case fatality rate as well as the asymptomatic rate in a rigorous way by factoring out the variabilities associated with the relevant studies. To accommodate the effects of missing asymptomatic infections on calculating the case fatality rate, we further perform a sensitivity analysis for the estimation of the case fatality rate. Our study provides a comprehensive evaluation of key measures of COVID-19 by taking into account of the heterogeneity and measurement error effects which are intrinsically associated with COVID-19 data. Our results offer sensible estimates of the clinical features of COVID-19 to enhance the understanding of the disease.

## 2 Method of Data Collection

### 2.1 Search Strategy and Selection Criteria

The third author (Y.Z.) conducted a literature screening for the articles published between January 24, 2020 and March 31, 2020 by using online databases, including PubMed, Web of Science, Google Scholar and the official websites of core scientific and biomedical journals including *Science, Nature, The Lancet, The New England Journal of Medicine*, and *The Journal of American Medical Association*, as well as some preprint platforms such as BioRxiv and MedRxir, with search terms specified as COVID-19, SARS-CoV-2, 2019-nCov, and novel coronavirus. Forty-three articles were found with the theme on the basic reproduction number, the incubation period, the percentage of asymptomatic cases, and the case fatality rate. Among those articles, 20 articles, described in Table 1, were identified by the first author (W.H.) to be included in the analysis, together with [24, 25] which were found on April 2. The inclusion criteria are the availability of both point estimates and 95% confidence intervals (or equivalently, standard deviations) for the basic transmission number, the average incubation time, the asymptomatic rate, or the case fatality rate.

### 2.2 Data Extraction and Analysis

Table 1 presents the summary information of the selected articles together with the descriptions of the data used in those articles. We extract the results for the basic reproduction number from [3, 4, 5, 6, 7, 9, 10] and the results for the average incubation time from [3, 4, 8, 12, 15]. The results from [20, 21, 22, 23, 24, 25] are extracted for estimation of the asymptomatic infection rate. The estimates for the case fatality rate together with their 95% confidence intervals are taken from [10, 11, 13, 14, 16, 17, 18]. In the articles [4, 6, 8], the reported 95% confidence intervals were asymmetric which we suspect were caused by employing a transformation (such as the exponential transformation) to the initial confidence intervals for the reparameterized effective size; for example, some authors may apply the logarithm to reparameterize the basic reproduction number or the average incubation time before performing the analysis. Using the inverse transformation, we convert the reported asymmetric confidence intervals and work out the associated standard deviations which are used in determining the weights for the meta-analysis.

**Table 1:**
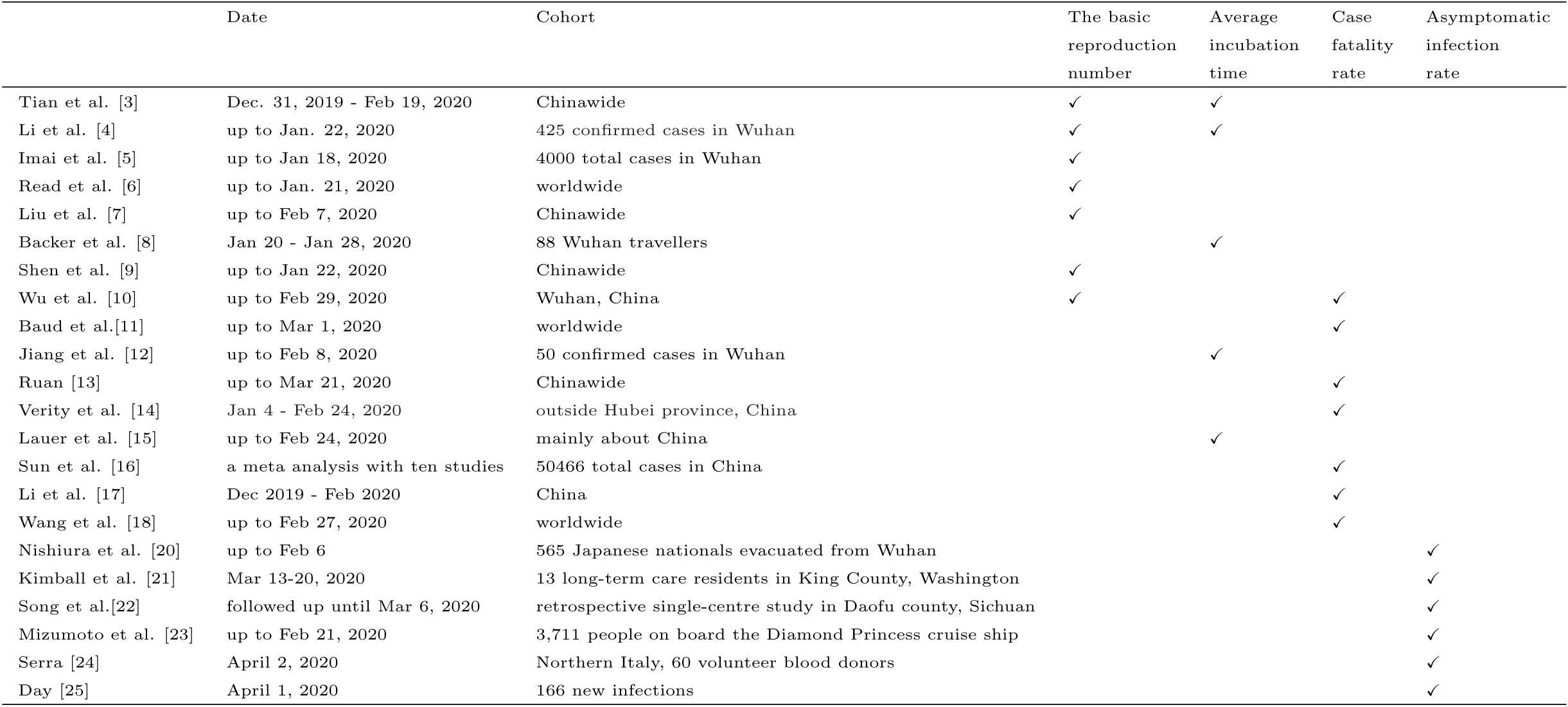
Descriptions of the articles included in the analysis.

## 3 Meta-Analysis

### 3.1 Method

As shown in the top panel of Figures 1–4, estimates of the basic reproduction number, the mean incubation time, the asymptomatic infection rate, and the case fatality rate are quite different from study to study. To obtain synthetic results, we perform a meta-analysis to aggregate the information from multiple studies with the same estimand (or *effect size* of interest) yet different features including the differences in the data collection, the sample size, and the conditions. Suppose *K* studies report an estimate and the associate standard deviation for an effect size of interest. For the *i*th study with *i* = 1, …, *K*, let *Y_i_* denote the *effect size* of interest and let 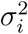 represent its associated variance estimate. In our analysis here, *Y_i_* is taken as the basic reproduction number, the average incubation time, the asymptomatic infection rate, and the case fatality rate, respectively. We calculate a weighted average of the results from those *K* studies under either the fixed effect model or the random effects model [26].

**Figure 1:**
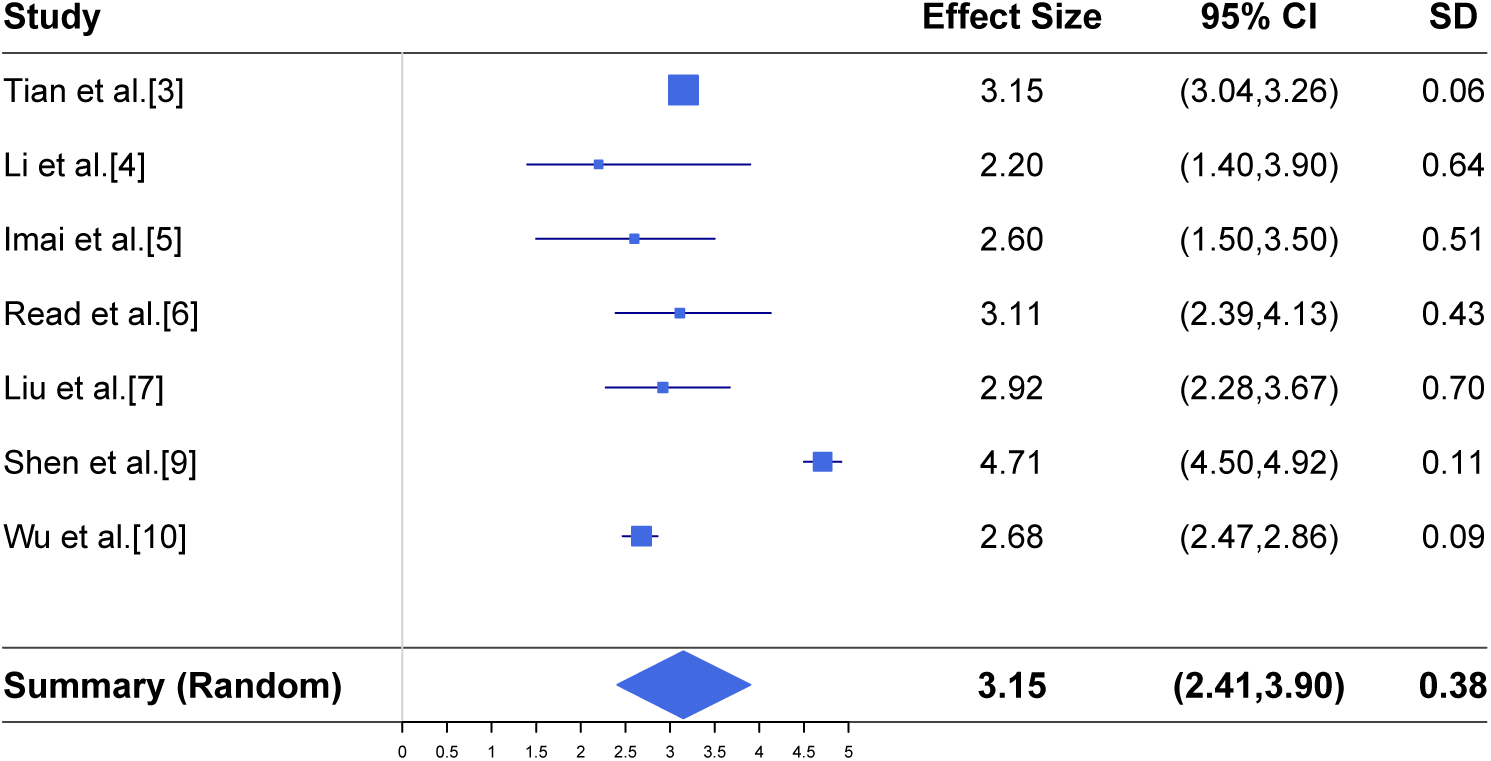
Meta-analysis of the basic reproduction number together with the reported results in seven different studies.

**Figure 2:**
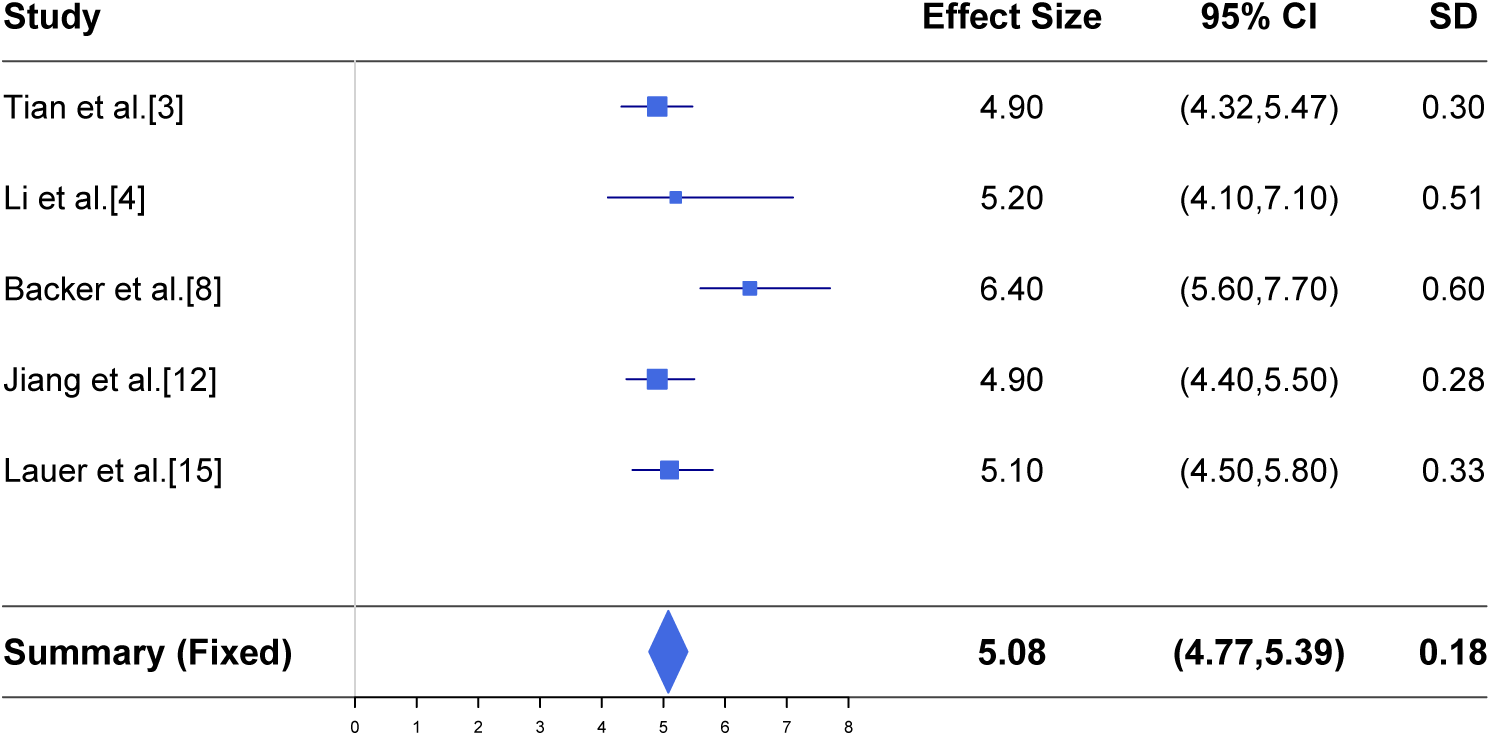
Meta-analysis of the mean incubation time (in day) together with the reported estimates in five different studies.

**Figure 3:**
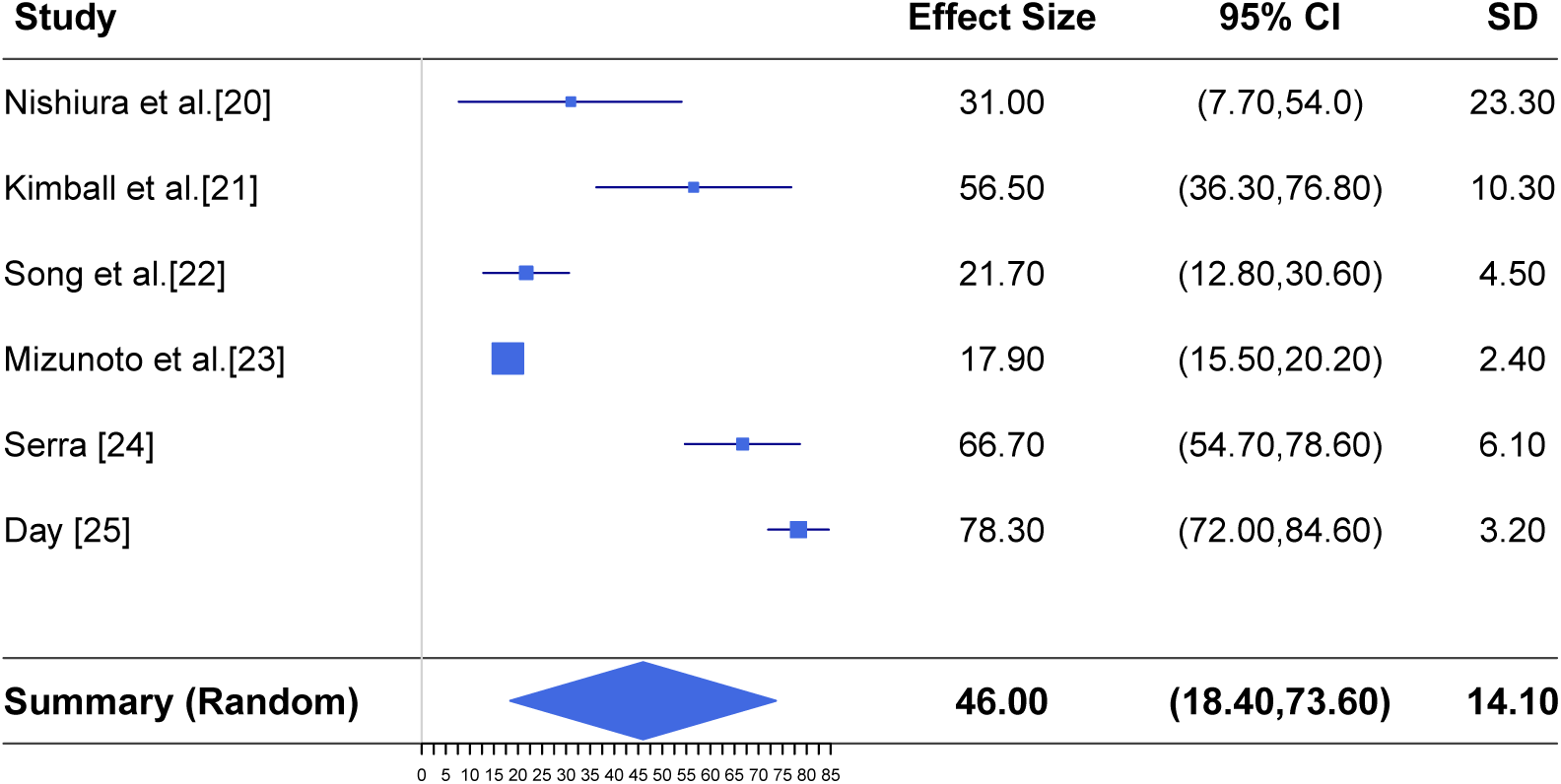
Meta-analysis of estimating the asymptomatic infection rate together with the reported results in six different studies.

**Figure 4:**
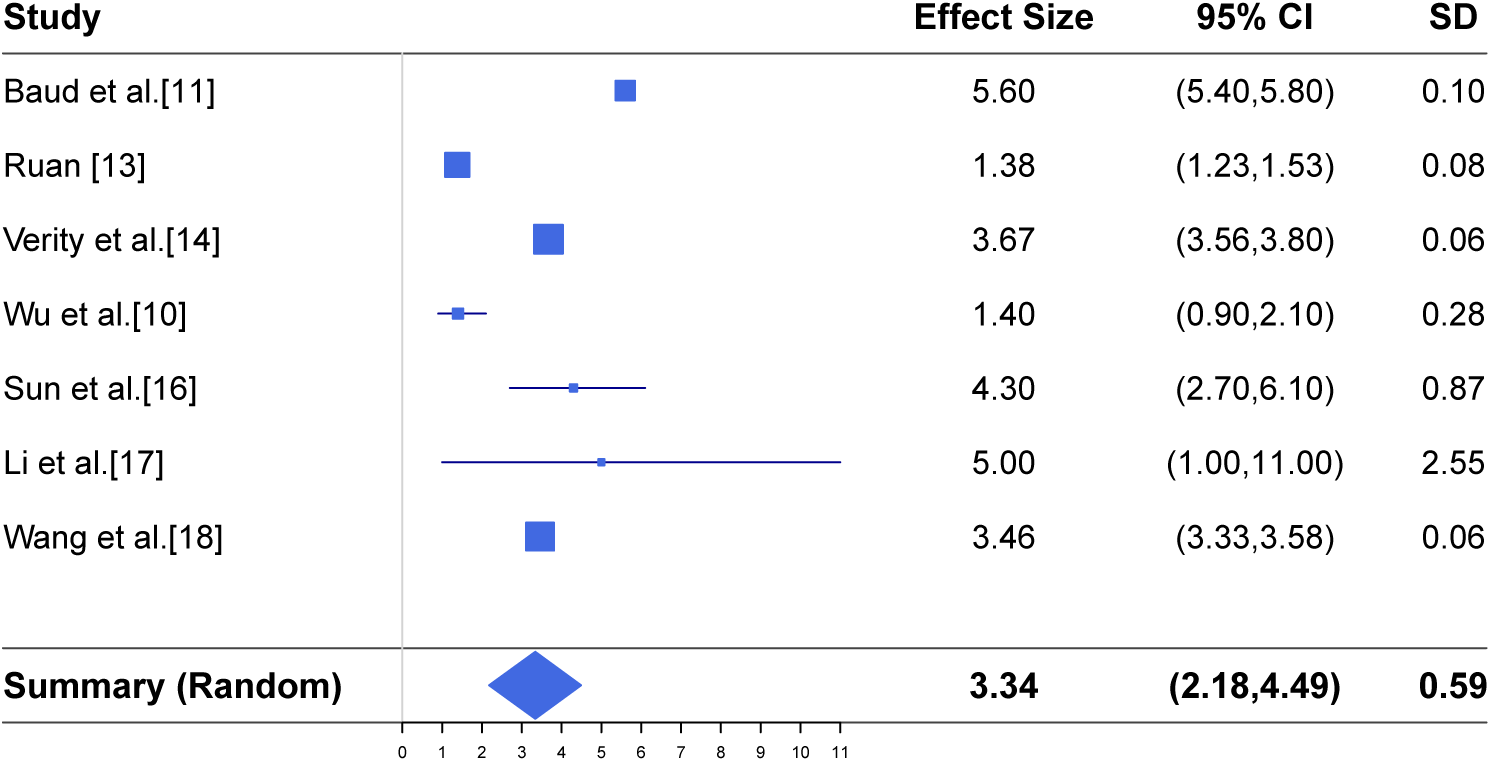
Meta-analysis of the case fatality rate (in percent) together with the results reported in seven different studies.

Under the fixed effect model, the meta mean effect size is given by

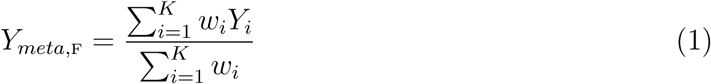

and the associated standard deviation is

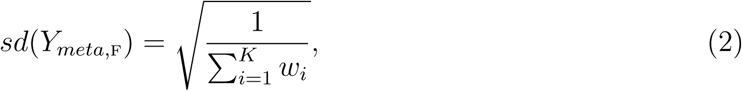

where 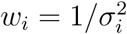 is the weight for the *i*th study.

With the random effects model, the meta mean effect size, denoted *Y_meta_*_,R_, and its standard deviation, denoted *sd*(*Y_meta_*_,R_), are determined by the same expression as (1) and (2) except for replacing the weight *w_i_* with a new weight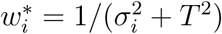, where

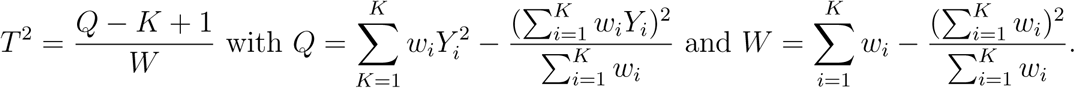

To determine whether the fixed effect model or the random effects model is suitable for the meta-analysis, we calculate the *I*^2^ index [28], defined as

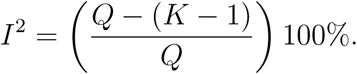

Consistent with [17], we take the fixed effect model if *I*^2^ < 50%, and the random effects model otherwise. In displaying the meta-analysis results, we use the R package *forestplot* [30].

### 3.2 Basic Reproduction Number

The top panel of Figure 1 shows the results for the basic production number reported in the seven studies. The *I*^2^ index for those studies is 97.8%, suggesting that the random effect model should be considered in conducting the meta-analysis. This result agrees with the perception that the basic reproduction number is time-dependent and varies from place to place.

The bottom panel of Figure 1 includes the meta-analysis results. The meta estimate of the basic reproduction number is 3.15, suggesting that a virus carrier may infect at least three individuals on average if preventive measures such as social distancing or quarantine are not applied to the public. It is noted that except for [3] and [7], other five studies listed in Figure 1 were based on the data for the earlier period of the outbreak where the lock down of Wuhan city has not been in effect yet. As more studies on the basic reproduction number become available for different places at different time periods, we can apply the same meta-analysis procedure to estimate the basic reproduction number to reflect its changes with the implementation of various measures to curb the virus spread in different regions.

### 3.3 Average Incubation Time

To estimate the average of the incubation time for infections, we study the results reported in the five articles summarized in the top panel of Figure 2. The *I*^2^ index is 28%, showing that the fixed effect model is suitable when conducting the meta-analysis. While the incubation time differs from patient to patient, varying between 1 and 14 days, as reported in [19], it is feasible to take their average time to be a fixed quantity.

The bottom panel of Figure 2 reports the meta-analysis results for the average incubation time of COVID-19. It shows that the mean incubation time is 5.08 days, with 95% confidence intervals being about 4.77 to 5.39 days. This estimated average incubation time is about 2 days shorter than the mean incubation time of 7 days announced by [19].

### 3.4 Asymptomatic Infection Rate

In the top panel of Figure 3 we display the estimates of the asymptomatic infection rate reported by [20, 21, 22, 23, 24, 25]. It is clear that those studies provided very different estimates of the asymptomatic infection rate, varying from 17.9% to 78.3%. Such a heterogeneity of the studies is confirmed by the I^2^ index which is 98%. Thus, we take the random effects model when conducting the meta-analysis. We report the results at the bottom panel of Figure 3. Our analysis suggests that the combined asymptomatic infection rate is 46% with the 95% confidence interval ranging from 18.4% to 73.6%.

### 3.5 Case Fatality Rate

Finally, we are interested in estimating the case fatality rate which measures how deadly COVID-19 is for the infected people. The meta-analysis results derived from seven studies available in the literature, shown in the top panel of Figure 4, are reported at the bottom panel of Figure 4, where we assume the random effect models because the *I*^2^ index is 99.5%. The estimated case fatality rate is 3.34%, slightly smaller than 3.4%, the estimate reported on March 3, 2020 by the WHO [2]. The 95% confidence interval suggests that the average case fatality rate can be as small as 2.18% and as large as 4.49%.

We comment that the *true* average case fatality rate is expected to be smaller than the estimate here, because the reported estimates of the case fatality rate in the literature were merely calculated as the ratio of the number of deaths from COVID-19 to the number of *reported* confirmed infected cases, where the number of *reported* confirmed infected cases is typically under-reported due to limited testing capacity and the exclusion of asymptomatic infections.

## 4 Sensitivity Analysis

To better understand what the true case fatality rate may be, we further conduct two sensitivity studies. In the first study, we repeat the meta-analysis of the case fatality rate in Section 3.5 by further including the results calculated from the data of the Princess Diamond cruise[29]. This analysis is driven by the consideration that the case fatality rate derived from the cohort of the cruise passengers is highly likely to be accurate, because the number of confirmed cases from the cruise is very likely to be close to the true number of infections. The bottom of Figure 5 reports the meta-analysis results obtained from the random effects model. With the inclusion of the results for the data of the Princess Diamond cruise, the weights of other seven studies become smaller than those in Figure 4, as shown by the size of the squares in Figures 4 and 5; and the estimate of the case fatality rate becomes smaller. The case fatality rate is estimated as 2.72% with the 95% confidence interval (1.29%, 4.16%).

**Figure 5:**
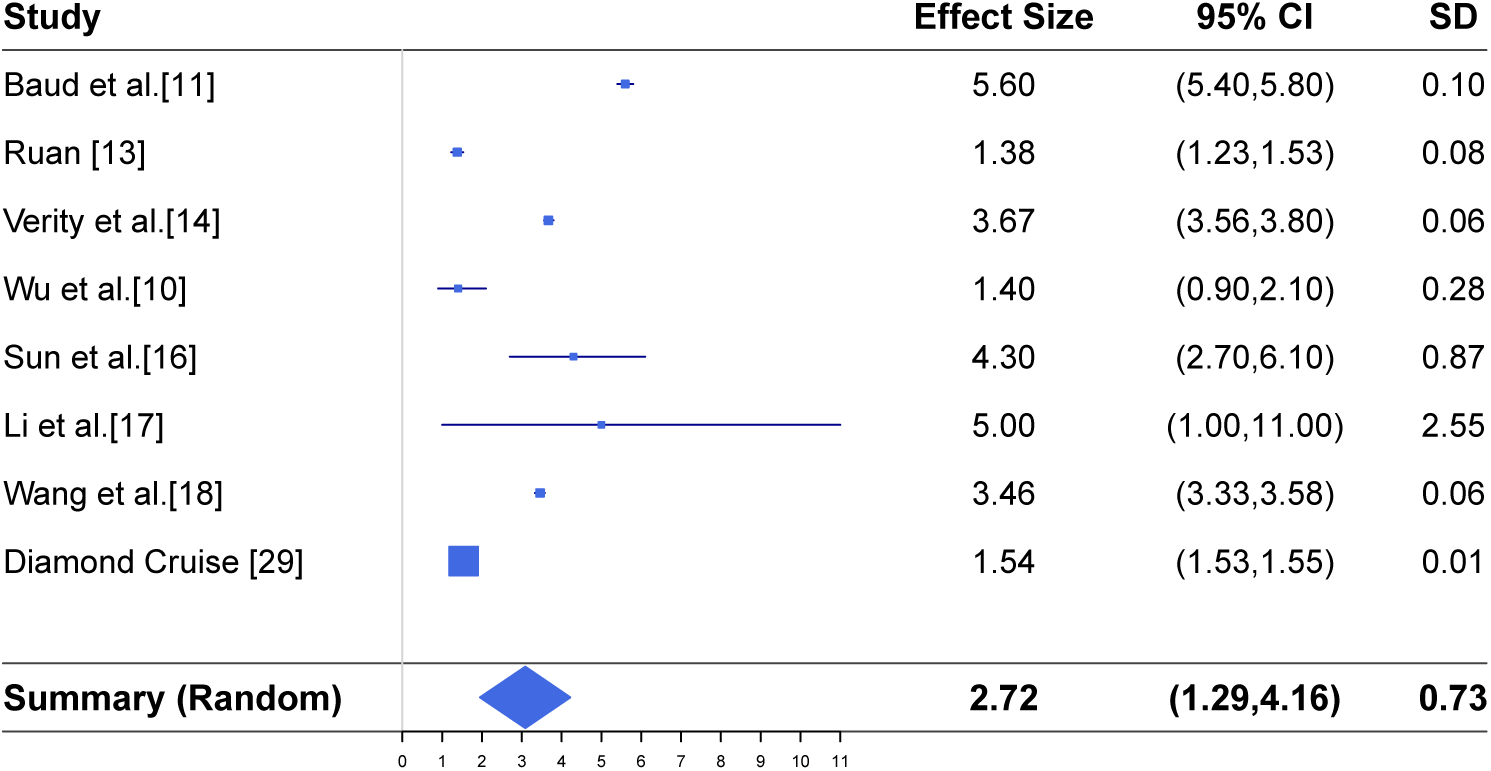
Meta-analysis of the case fatality rate (in percent) together with the results reported in seven different studies and the results from the Princess Diamond cruise.

In our second sensitivity analysis, we revise the results in Figure 4 by incorporating the information of asymptomatic cases. To see the adjustment, we let *D* represent the number of deaths caused from COVID-19. Let *C*_R_ denote the number of reported infected cases of COVID-19, let *C*_A_ stand for the number of the SARS-Cov-2 carriers who are asymptomatic, and let *C* be the total number of infected cases with the virus. Let *r*_A_ = *C*_A_/*C* be the ratio of asymptomatic infections to the true number of infections.

Let *p*_R_ = *D*/*C*_R_ be the *reported* case fatality rate and let *p*_T_ = *D*/*C* be the *true* case fatality rate. If we assume that *C* = *C*_R_ + *C*_A_, then the *reported* case fatality rate and the *true* case fatality differ by the factor 1 − *r*_A_:

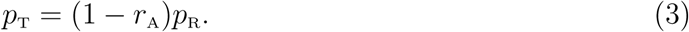

Estimates of the case fatality rate that have been reported in the current literature are merely directed to *p*_R_ rather than *p*_T_.

To sensibly estimate the *true* case fatality, we use (3) to adjust the reported results of the seven studies listed at the top panel of Figure 4. Specifically, we may multiply the factor 1 − *r*_A_ with an estimate for the reported rate *p*_R_ as well as its standard deviation for each study and then run a meta-analysis. However, the exact value of the asymptomatic infection rate is unavailable, and we only have its estimates from various studies displayed at the top panel of Figure 3. To assess how the uncertainty of not knowing the true value of *r*_A_, we use two ways to set a value for *r*_A_ to modify the reported fatality rates for the studies listed at the top panel of Figure 4 for running a new meta-analysis.

First, taking *r*_A_ as one of seven reported estimates listed at the top panel of Figure 3, we modify the reported results provided by each study listed at the top panel of Figure 4 using (3), and report the meta analysis results at the top panels of Figure 6. In the second analysis, we take *r*_A_ as the synthesized estimate reported in Figure 3, i.e, *r*_A_ is set as 46%, and then run the meta-analysis for these adjusted case fatality rates under the random effects model. We report the results at the bottom panel of Figure 6, which shows that the estimate of the case fatality rate is 1.8% with the 95% confidence interval ranging from 1.18% to 2.43%.

**Figure 6:**
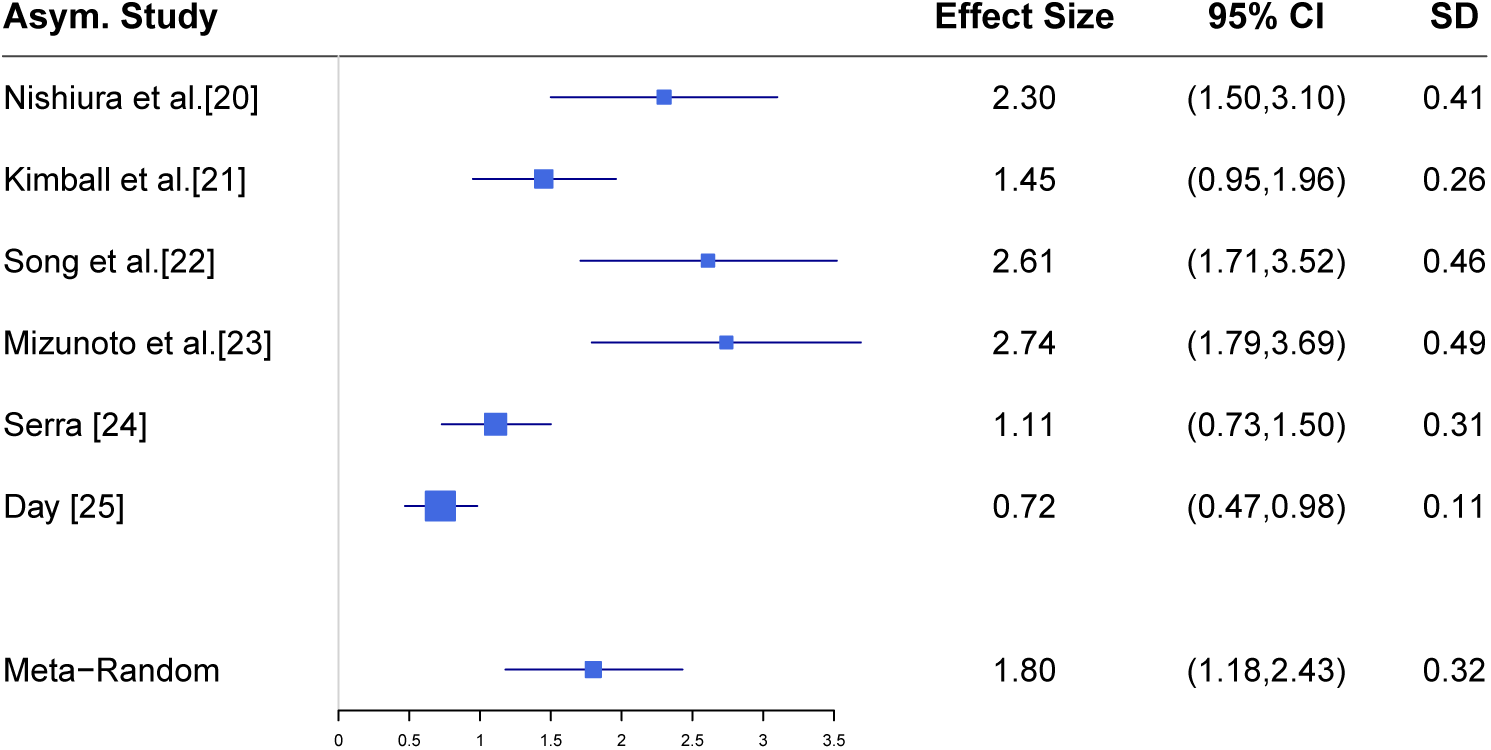
Sensitivity analysis of estimating the case fatality rate with different asymptomatic rates accounted for.

## 5 Conclusions and Discussion

We carry out a meta-analysis and sensitivity study for estimating the basic reproduction number, the average incubation time, the asymptomatic infection rate, and the case fatality rate for COVID-19. Examining the published results between January 24, 2020 and March 31, 2020, our study aggregates different results reported in the literature and provides synthetic estimates by addressing the heterogeneity present in different studies. Our study shows that the basic reproduction number is estimated to be 3.15 with the 95% confidence interval (2.41, 3.90) and the average incubation time is 5.08 days with the 95% confidence interval ranging from 4.77 days to 5.39 days. The asymptomatic infection rate is estimated to be 46% with the 95% confidence interval (18.48%, 73.60%). While multiple studies reported estimates of the case fatality rate, those estimates are typically higher than the true case fatality rate under the same conditions, which is attributed to the fact that a good portion of asymptomatic infections are not counted when estimating the case fatality rate. Our sensitivity study addresses this important issue and makes an adjustment to provide a sensible estimate of the case fatality rate. Compared to the estimated 3.34% case fatality rate obtained from the meta-analysis, our sensitivity study estimates the case fatality rate to be 1.8% with 95% confidence interval (1.18%, 2.43%) where asymptomatic infections are accounted for.

Our studies reveal sensible estimates for the important quantities of COVID-19 by accommodating discrepancy effects associated with different studies such as the variability of the data collected from different populations at different time periods. With the evolution of the pandemic, the basic production number can greatly reduce as a result of the implementation of active measures to mitigate the virus spread. The estimation of the case fatality rate may be closer to the true case fatality rate because of the increase of the test capacity; more infected cases may be detected so the reported number of infections would be closer to the true number of COVID-19 carriers. Our results are useful in enhancing the knowledge of COVID-19. Though we focus on evaluating the basic reproduction number, the average incubation time, the asymptomatic infection rate, and the case fatality rate, other features, such as the time from symptom onset to hospitalization or to death and the morbidity rate of the disease, are also important and they are worth being estimated in a sensible way.

The present investigations have limitations. Not all published results for the four measures are included in our study; we do not include those manuscripts which reported merely a point estimate without the associated standard deviation or a 95% confidence intervals, because they do not allow us to decide a proper weight for the inclusion of the result. While reporting a single estimate of the average incubation time and the case fatality rate gives us an easy way to assess the impact of COVID-19, such measures marginalize the effects from the associated factors such as the disease severity, the patient’s medical conditions, and age. With more studies available for categorizing the case fatality rate or the incubation time, it is useful to apply the meta-analysis to estimate those measures by stratifying the population based on the demographic and clinical characteristics. When data at the individual level are available, better estimates of key features for COVID-19 can be obtained and the pandemic trend can be more reasonably projected using statistical regression models.

## Data Availability

This research is a literature review and meta-analysis. Thus, no new data are involved.

## Acknowledgements

The research is partially supported by fundings from the Natural Sciences and Engineering Research Council of Canada (NSERC). Yi is Canada Research Chair in Data Science (Tier 1). Her research was undertaken, in part, thanks to funding from the Canada Research Chairs Program.

## Author Contributions

WH identified the articles that were screened by the third author, extracted the results from individual studies, conducted all the data analyses, and prepared the initial draft. GY discussed the analysis methods with WH and wrote the manuscript. YZ searched the literature, provided the candidate articles to WH for further examination, and help formatting the article according to the journal template.

## References

[1] World Health Organization (2020) COVID-19 Situation Reports. https://www.who.int/emergencies/diseases/novel-coronavirus-2019/situation-reports

[2] World Health Organization (2020) WHO Director-General’s opening remarks at the media briefing on COVID-19 - 3 March 2020. March 3, 2020

[3] Tian H, Liu Y, Li Y, Wu C-H, Chen B, Kraemer MUG, Li B, Cai J, et al. (2020) An investigation of transmission control measures during the first 50 days of the COVID-19 epidemic in China Science. Nature, 2020, online on Mar. 31, 2020.

[4] Li Q, Guan X, Wu P, Wang X, Zhou L, Tong Y, et al. (2020) Early transmission dynamics in Wuhan, China, of novel coronavirus-infected pneumonia. New England Journal of Medicine, 2020. https://doi.org/10.1056/NEJMoa2001316.

[5] Imai N, Dorigatti I, Cori A, Riley S, and Ferguson NM (2020) Report 3: Transmissibility of 2019-nCoV. Online at https://www.imperial.ac.uk/mrc-global-infectious-diseaseanalysis/covid-19/report-3-transmissibility-of-covid-19/.

[6] Read JM, Bridgen JRE, Cummings DAT, Ho A and Jewell CP (2020) Novel coronavirus 2019-nCoV: early estimation of epidemiological parameters and epidemic predictions. *medRxiv*. DOI: https://doi.org/10.1101/2020.01.23.20018549v2.

[7] Liu T, Hu J, Kang M, Lin L, Zhong H, Xiao J, et al. (2020) Time-varying transmission dynamics of Novel Coronavirus Pneumonia in China. *BioRxiv*, 2020. DOI: https://doi.org/10.1101/2020.01.25.919787.

[8] Backer JA, Klinkenberg D and Wallinga J (2020) Incubation period of 2019 novel coronavirus (2019-nCoV) infections among travellers from Wuhan, China, 20–28 January 2020. Eurosurveillance. https://doi.org/10.2807/1560-7917.ES.2020.25.5.2000062.

[9] Shen M, Peng Z, Xiao Y and Zhang L (2020) Modelling the epidemic trend of the 2019 novel coronavirus outbreak in China. bioRxiv. https://doi.org/10.1101/2020.01.23.916726.

[10] Wu JT, Leung K and Leung GM (2020) Nowcasting and forecasting the potential domestic and international spread of the 2019-nCoV outbreak originating in Wuhan, China: a modelling study. Lancet, 395, 689–697.

[11] Baud D, Qi X, Nielsen-Saines K, Musso D, Pomar L and Favre G (2020) Real estimates of mortality following COVID-19 infection. Lancet Infectious Diseases. https://doi.org/10.1016/S1473-3099(20)30195-X.

[12] Jiang X, Rayner S and Luo MH (2020) Does SARS-CoV-2 has a longer incubation period than SARS and MERS? Journal of Medical Virology, 92, 476–478.

[13] Ruan S (2020) Likelihood of survival of coronavirus disease 2019. Lancet Infectious Diseases. Online on March 30, 2020, https://doi.org/10.1016/S1473-3099(20)30257-7.

[14] Verity R, Okell LC, Dorigatti I, Winskill P, Whittaker C, Imai N, et al. (2020) Estimates of the severity of coronavirus disease 2019: a model-based analysis. Lancet Infectious Diseases, Online on March 30, 2020. https://doi.org/10.1016/S1473-3099(20)30243-7.

[15] Lauer SA, Grantz KH, Bi Q, Jones FK, Zheng Q, Meredith HR, Azman AS, Reich NG, and Lessler J (2020) The Incubation Period of Coronavirus Disease 2019 (COVID19) From Publicly Reported Confirmed Cases: Estimation and Application. Annals of Internal Medicine. DOI:10.7326/M20-0504.

[16] Sun P, Qie S, Liu Z, Ren J, Li K and Xi J (2020) Clinical characteristics of hospitalized patients with SARS-CoV-2 infection: A single arm meta-analysis. Journal of Medical Virology. DOI: 10.1002/jmv.25735.

[17] Li L, Huang T, Wang Y, Wang Z, Liang Y, Huang T, Zhang H, Sun W and Wang Y (2020) COVID-19 patients’ clinical characteristics, discharge rate, and fatality rate of meta-analysis. Journal of Medical Virology. DOI: 10.1002/jmv.25757.

[18] Wang Y, Wang Y, Chen Y and Qin Q (2020) Unique epidemiological and clinical features of the emerging 2019 novel coronavirus pneumonia (COVID-19) implicate special control measures. Journal of Medical Virology. DOI: 10.1002/jmv.25748.

[19] National Health Commission of People’s Republic of China (2020) Prevent guideline of 2019-nCoV. http://www.nhc.gov.cn/xcs/yqfkdt/202001/bc661e49b5bc487dba182f5c49ac445b.shtml (in Chinese). Accessed April 10, 2020.

[20] Nishiura H, Kobayashi T, Suzuki A, et al. (2020) Estimation of the asymptomatic ratio of novel coronavirus infections (COVID-19). International Journal of Infectious Diseases. DOI:10.1016/j.ijid.2020.03.020

[21] Kimball A, Hatfield KM, Arons M, et al. (2020) Asymptomatic and Presymptomatic SARS-CoV-2 Infections in Residents of a Long-Term Care Skilled Nursing Facility – King County, Washington, March 2020. MMWR Morbidity Mortality Weekly Report, 69, 377–381.

[22] Song H, Xiao J, Qiu J, et al. (2020) A considerable proportion of individuals with asymptomatic SARS-CoV-2 infection in Tibetan population. *MedRxiv*. DOI:10.1101/2020.03.27.20043836

[23] Mizumoto K, Kagaya K, Zarebski A, et al. (2020) Estimating the asymptomatic proportion of coronavirus disease 2019 (COVID-19) cases on board the Diamond Princess cruise ship, Yokohama, Japan, 2020. Eurosurveillance, 25. DOI:10.2807/1560-7917.ES.2020.25.10.2000180

[24] Serra M (2020) Coronavirus, Castiglione d’Adda è un caso di studio: ‘Il 70% dei donatori di sangue è positivo’. Top News, Lastampa, Italy. April 2, 2020. https://www.lastampa.it/topnews/primo-piano/2020/04/02/news/coronaviruscastiglione-d-adda-e-un-caso-di-studio-il-70-dei-donatori-di-sangue-e-positivo1.38666481

[25] Day M (2020) Covid-19: four fifths of cases are asymptomatic, China figures indicate. BMJ 369:m1375 DOI: 10.1136/bmj.m1375 (Published 2 April 2020)

[26] Borenstein M, Hedges LV, Higgins JPT and Rothstein HR (2009). Introduction to Meta-Analysis. Wiley.

[27] Gordon M and Lumley T (2019) R package “Forestplot”. https://cran.r-project.org/web/packages/forestplot/forestplot.pdf

[28] Higgins JPT and Thompson SG (2002) Quantifying heterogeneity in a meta-analysis. Statistics in Medicine, 21, 1539–1558.

[29] John Hopkins University (2020) 2019 Novel Coronavirus COVID-19 (2019-nCoV) Data Repository by Johns Hopkins CSSE. Access on April 1, 2020.

[30] Gordon M and Lumley T (2019) R package “Forestplot”. https://cran.r-project.org/web/packages/forestplot/forestplot.pdf

